# Heart-retina time analysis using electrocardiogram-coupled time-resolved dynamic optical coherence tomography

**DOI:** 10.1101/2024.07.15.24310387

**Authors:** Philippe Valmaggia, Julia Wolleb, Florentin Bieder, Hendrik P.N. Scholl, Philippe C. Cattin, Peter M. Maloca

## Abstract

The eye and the heart are two closely interlinked organs, and many diseases affecting the cardiovascular system manifest in the eye. To contribute to the understanding of blood flow propagation towards the retina, we developed a method to acquire electrocardiogram (ECG) coupled time-resolved dynamic optical coherence tomography (OCT) images. This method allows for continuous synchronised monitoring of the cardiac cycle and retinal blood flow dynamics. The dynamic OCT measurements were used to calculate time-resolved blood flow profiles using fringe washout analysis. The relative fringe washout was computed to generate the flow velocity profiles within arterioles at the optic nerve head rim. We found that the blood column between the heart and the retina propagates within one cardiac cycle, denoting the arrival time as the heart-retina time (HRT). In a group of healthy subjects, the HRT was 144 ± 19 ms (mean ± SD). The HRT could provide a novel potential biomarker for cardiovascular health in direct relation to retinal perfusion.

## Introduction

The eye and the heart are closely connected ^1–3^. The cardiovascular system plays a crucial role in delivering blood to the eye. The blood travels through a sequence of arteries, including the carotid, ophthalmic, and central retinal arteries, until it reaches the retina. The fine bed of capillary structures guarantees the delivery of nutrients at the retina, which is essential for maintaining the visual function. Perturbances of blood flow towards the eye can cause ischemic occlusions or haemorrhages, ultimately leading to vision loss ^4–7^. The heart’s function can be analysed with the electrocardiogram (ECG), precisely measuring the cardiac rhythm and anomalies ^8^. Analysing the cardiac cycle enables us to detect the moment of contraction of the ventricles, which leads to the blood being pumped towards the corporal circulation.

The eye, specifically the retina, can be imaged with optical coherence tomography (OCT) ^9^. OCT allows for a non-invasive assessment of ocular structures and generates high-resolution images on a micrometre scale ^10^. Recent developments in OCT have enabled us to measure OCT dynamically, even with devices approved for routine clinical use ^11^. These dynamic OCT acquisitions allow the estimation of dynamic flow profiles within vessels of the retinal vasculature and were suggested to represent a propagation of the cardiac cycle pulsation ^11^. These time-resolved dynamic blood flow profiles surpass certain limitations of optical coherence tomography angiography (OCTA) providing qualitative perfusion maps, variable inter-scan time analysis (VISTA) providing quantitative single-velocity (per vessel) perfusion maps, or laser speckle flowgraphy (LSFG) providing en-face flow profiles ^12–14^. The time-resolved dynamic OCT acquisitions allow a depth-resolved flow profile analysis with a temporal resolution as fast as 101 Hz per B-scan for an acquisition with 125 kHz per A-scan on B-scans with 1024 A-scans.

However, the propagation of the cardiac pulse wave is poorly understood because the cardiac and retinal acquisitions are not synchronised. Coupling devices could provide additional information about potential mismatches between the cardiac cycle pulse waves and the pulse waves recorded at the retinal level. To this end, we investigated the feasibility of linked acquisitions by coupling an ECG to a time-resolved OCT device. The importance of timestamp matching must be emphasised here, which is why it was decided to acquire the ECG information with an open-source device capable of recording the ECG signal with microsecond precision. The time-resolved OCT images have integrated timestamps, which we can use to match the two time series. Both clocks were synchronised using a network time protocol (NTP) server.

We aimed to generate a new option to analyse information regarding the propagation of cardiac pulsations towards the eye by linking the information of the ECG to the time-resolved OCT. Knowledge about the propagation velocity of the cardiac pulse wave could be helpful in finding potential turbulences in the pathway and give a general overview of cardiovascular health, and specifically, risks for cardiovascular disturbances manifesting in the retina. We introduce the heart-retina time (HRT), which represents the time the blood column needs to propagate from the heart to the eye. We further aim to present with this work the feasibility of ECG-coupled time-resolved OCT acquisitions, the correlation between the cardiac cycle and retinal pulse waves and the potential of the HRT as a biomarker.

## Methods

### Study design and patient population

This was a cross-sectional observational cohort study in healthy subjects. The inclusion criteria contained an age of at least 18 years and no history of vascular diseases or ophthalmic surgery. Further, the participants did not smoke or drink coffee six hours before the measurements to prevent possible effects on the retinal microvasculature ^15^. Each subject was informed about the study and written informed consent was obtained. The study was approved by the Ethics Committee Northwestern and Central Switzerland (EKNZ_2021-02360) and performed in accordance with the declaration of Helsinki.

### Hardware description

The OCT device used for this study was a Spectralis (Heidelberg Engineering GmbH, Heidelberg, Germany). The device allowed for a modification of the scan patterns and the integration time with an investigational acquisition module ^16,17^. The ECG device used for this study was the open-source vitals monitor Healthy Pi v4.4 with a 3-lead ECG (Protocentral, Bangalore, India). The Healthy Pi v4.4, based on an ESP32 microprocessor, had the capability to record ECGs with a temporal frequency of 125 Hz ^18^. The Arduino code was modified to include timestamps with a microsecond precision into the same data packet as the ECG samples ^19^. Power supply and data transfer were done with a high-speed micro-USB 2.0 cable and connected to the same computer as the Spectralis. The programming of the ECG was performed with the Arduino language in IDE V 1.8.13 and a graphical user interface was generated with Processing V 3.5.4.

The timestamps of the OCT device were generated via the computer clocks whereas the timestamps for the ECG acquisitions were generated via the ESP32 microprocessor clock. The clocks were synchronised to the NTP server ntp11.metas.ch of the Swiss Federal Office of Metrology and Accreditation ^20^. The ESP32 microprocessor was synchronised via WIFI and the synchronisation of the OCT timestamps was performed via a LAN cable connection. A schematic overview of the acquisition setup can be found in Figure 1.

**Fig. 1.**
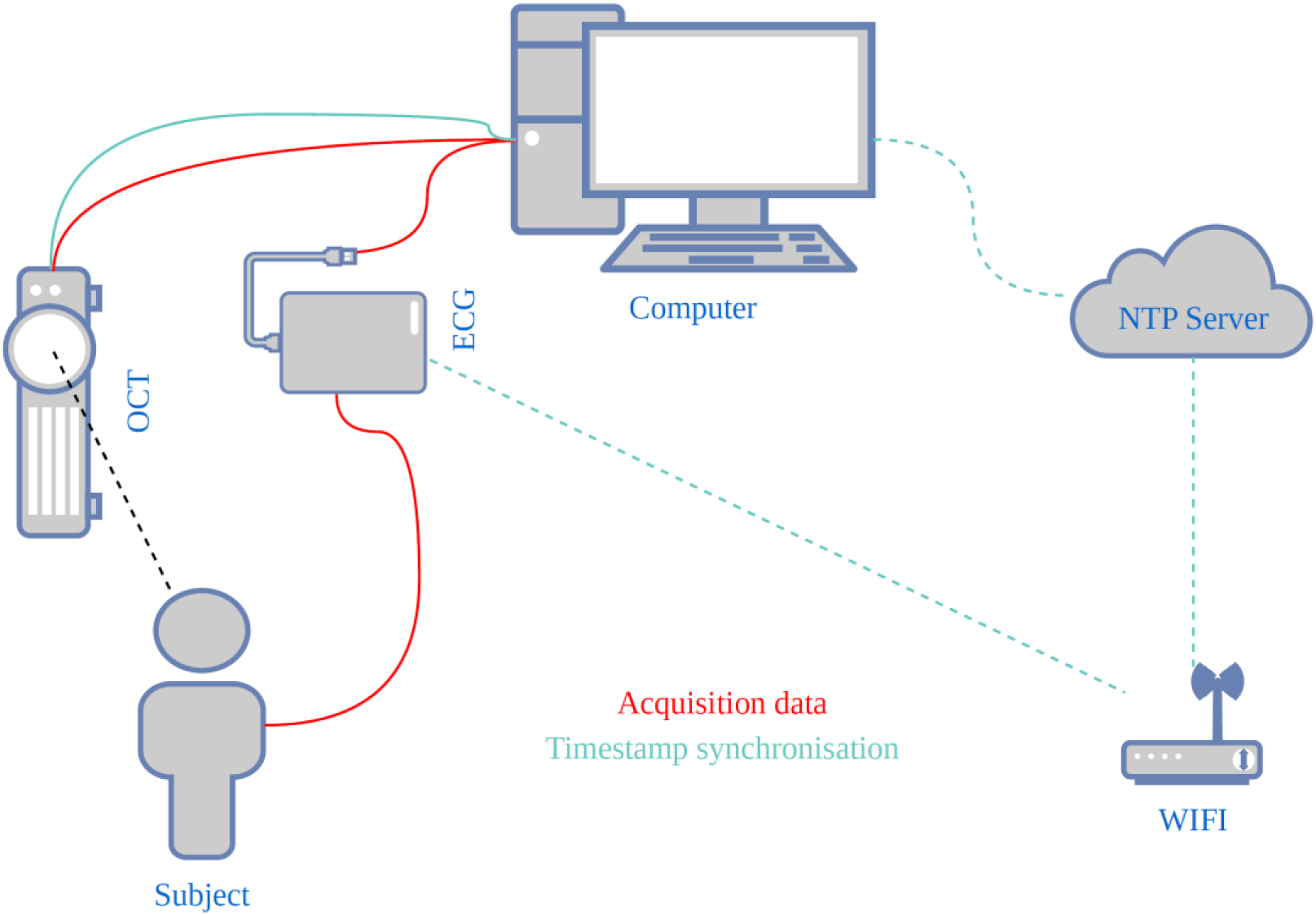
Electrocardiogram-coupled optical coherence tomography. Time-resolved dynamic OCT and ECG data are recorded from a subject synchronously. The data is transferred to a central computer. The timestamps of the ECG and the OCT are synchronised via a Network Time Protocol (NTP) server. The OCT timestamps are synchronised via a direct connection to a stratum 1 NTP server. The timestamps of the ECG are synchronised with the same NTP server via timestamp synchronisation request via WIFI.

### Data acquisition

The demographic characteristics of the study subjects (age, sex, height) were obtained from the study subjects before the measurements. An example OCT volume was acquired to familiarise the study subjects with the OCT device and the specific fixation target. Following this initial test, the patients were connected to the ECG device and seated in front of the OCT. Timestamp synchronisation was performed and tested before the acquisitions started.

Three-lead ECG acquisitions were started, and data were continuously recorded to a .csv file, including the timestamps with a microseconds precision and the ECG voltage in arbitrary units generated by the HealthyPi (HP value). The OCT acquisition module was then initiated, and the patient asked to remain calm and fixate on the blue cross target provided by the manufacturer. An equal illumination of the scanning laser ophthalmoscopy (SLO) in all image areas was verified and the acquisition location of the B-Scan was steered to the desired position. Continuous B-scans were then acquired at the superior and inferior optic nerve head rims of both eyes. The nominal acquisition line-scan rate was set to 20 or 85 kHz, corresponding to an integration time of 44.8 or 11.2 µs, respectively. The B-scans were continuously acquired for approximately 7 seconds with a scan pattern of 1024 × 496 pixels on a field-of-view (FOV) of 10°. In case of blinks or bad signal quality, the recorded data were not used for further analysis.

### Data processing

The ECG data were resampled to fixed interval timestamps at the 125 Hz acquisition rate with linear interpolation for missing ECG values. Following this, the R-Peaks of the ECG were extracted using the “EngZee” algorithm with an implementation in Python ^21,22^. The binary R-Peak signals were then smoothed using a Gaussian filter (σ **=** 1.25) and the values afterwards normalised between 0 and 1. The heart rate was then given by the R-Peaks.

The OCT data were exported in RAW format (.vol). Each of the B-scans was matched with a timestamp with milliseconds precision. The timestamps were exported with the corresponding image included in the .vol file header. Pixel intensities in the .vol format are stored in normalised values between 0 and 1. For visualisation purposes, these pixel intensities were transformed for each as 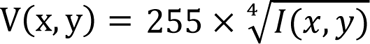, where V(x,y) denotes the pixel intensity for visualisation at coordinates (x,y) and I(x,y) denotes the normalised original intensity.

As a first step after intensity transformation, the B-scans were registered with a rigid transformation using a pyramid processing framework ^23^. The framework was modified and an average of the first ten images of each time series image stack was taken as reference image for the registration of the B-scans on each other. The annotation of the vessel centres was then manually performed. For this, the registered volumes were loaded as 3D image stacks into 3D Slicer V4.11. The arterial centres were manually annotated as landmarks in each single B-Scan. The vessel centres were identified by 1) identifying the horizontal centres point by identifying the shadow behind the vessel, and 2) Identifying the anterior-posterior centres by identifying the centre between the hyperreflective vessel walls ^24^.

Subareas surrounding the arteriole centres of 7×7 pixels (corresponding to ∼20×27 μm) were then extracted along the time series. Inside these subareas, the signal-to-noise ratio SNR was calculated by dividing the average signal intensity of the subarea divided by the noise level of the B-scan. Assuming shot-noise limited detection, the B-scan noise level was approximated by dividing the maximum intensity extracted from the raw data by the Heidelberg Quality score. The SNR for the subarea of each B-Scan was calculated as:

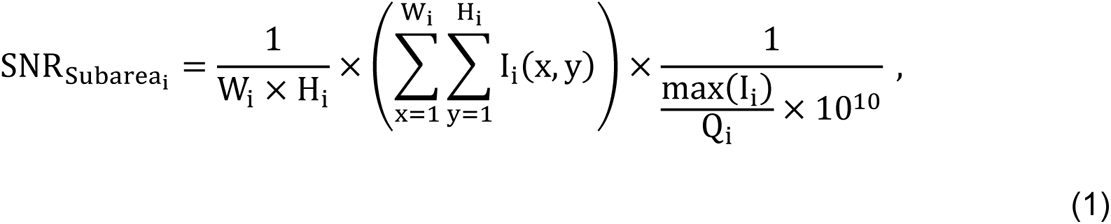

where i denotes the index of the B-scan, I(x,y) denotes the intensity at the coordinates (x, y).The width and height of the subarea are denoted as w and h. 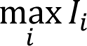 denotes the maximum intensity obtained from the metadata of the B-scan, and Q denotes the Heidelberg Quality score in decibel (dB). Each i-th B-scan was matched to a timestamp with ms precision, which was also stored in the metadata.

The SNR of each subarea was then compared with a reference SNR to calculate the drop in SNR (SNR_Drop_). As reference level, the peak SNR on a pixel level was chosen. The SNR_Drop_ was used to calculate relative flow velocity profiles according to^11,25^:

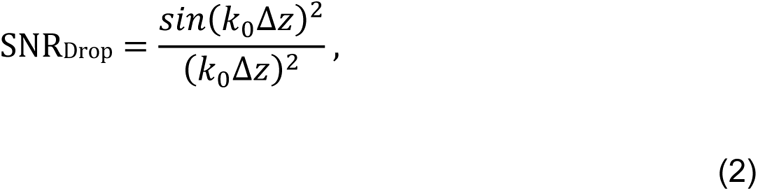

where k_0_ = 2 * π / λ and Δz = n * v * τ. The equation was solved numerically for v, the velocity of the axial component of the motion. In these equations, k_0_ denotes the central wavenumber, λ is the central wavelength of the light source, Δz corresponds to the displacement in the z-axis, n is the eye’s refractive index, and τ denotes the integration time. Multiple possible solutions exist, and every numerical solution calculated for each B-scan was stored as a potential flow velocity.

The pulse wave analyses for the correlation with the cardiac cycle were performed with the envelope function of the maximal numerical solution of Eq. (2). Local maxima of the first derivative of the flow profile were taken to identify the sharpest rises of the blood flow velocity, representing the arrival time of the pulse propagation at the retina. The local maxima were identified within the time series bins correlating to the heart rate, which was identified by the ECG. These local maxima were then processed in the same way as the R-Peaks: They were binarised along the time axis as single time points, and the Gaussian filter (σ **=** 1.25) was applied, followed by a [0,1] normalisation.

The timelines of the ECG R-Peaks and the pulse arrival times of the time-resolved OCT data were then compared with cross-correlations. This was used to calculate the lag between the two time series, where the OCT time series served as the baseline for the cross-correlation. The ECG data was extracted from 10 seconds prior to the first B-scan of the time-resolved OCT data until 1 second after the last B-scan. The lag values were then sorted according to the argmax of the cross-correlation values to identify the most probable offset between the moment of the contraction of the heart ventricles (ECG R-Peaks) and the arrival at the retina (sharpest rise in OCT blood flow velocity profile).

The HRT was then denominated for the time needed for the arrival at the retina. This time corresponds to the time the impulse to the blood column needs to arrive from the contraction of the left cardiac ventricle, as identified by the R-Peak at the retina. This pulse propagation times aim to reveal information about the rigidity of the vessels. Hence, information about the functioning of the cardiovascular system can be obtained by combining recordings of the electrical heart activity with an ECG and imaging of the retina with structural OCT.

The timestamp synchronisation of the OCT and the ECG was performed via timestamp synchronisation with the NTP server, including an offset correction for asymmetric network synchronisation between both devices. Several synchronisations have been completed within the same network to assess the reliability. The programming tasks for image analysis were carried out using Linux shell scripts, Python 3.9 (Python Software Foundation, Wilmington, USA), and R V4.2.2 (developed by the Foundation for Statistical Computing, Vienna, Austria). For data visualisation, Python 3.9, R V4.2.2, and 3DSlicer V4.11 were used ^26^.

### Statistical analysis

The statistical analysis focused on different aspects of the data. First, we analysed the flow velocity profiles, including the acquisition characteristics, the calculated velocities and the SNR. The correlation with the ECG data was then analysed qualitatively by plotting the time series and quantitatively with cross-correlations. The reproducibility of the HRT was assessed for intra-subject, inter-subject, intra-exam, and intra-vessel variability. These variabilities were analysed with the coefficient of variation (CoV). Descriptive values in this manuscript are presented as mean ± standard deviation (SD) or median [interquartile-range (IQR)].

## Results

### Feasibility of coupled ECG and time-resolved OCT acquisitions

The synchronous ECG and OCT data acquisition, including timestamp synchronisation, was feasible and well-tolerated in all subjects. Two subjects were female, and three were male. The subjects were 24 to 32 years old and healthy. In total, 90 arterioles were analysed in 70 acquisitions at the optic nerve head. In right eye acquisitions (n=47), 25 arterioles have been analysed at the superior part of the optic nerve head and 22 at the inferior part. In left eye acquisitions, 23 arterioles have been analysed at the superior part and 20 at the inferior part. 54 arterioles from 39 acquisitions have been acquired at a nominal A-scan rate of 20 kHz, and 36 arterioles from 31 acquisitions have been acquired at a nominal A-scan rate of 85 kHz. From these acquisitions, a total of 26’100 arteriole centres have been manually annotated in all B-scans combined. Further information about the acquisition parameters can be found in Table 1, including an overview of the calculated velocity flow profiles.

**Table 1.** Acquisition overview per nominal A-scan rate.

|  | **20 kHz, n = 54** | **85 kHz, n = 36** |
| --- | --- | --- |
| **Exam Duration [ms]** | 7144<br>(6908, 7480) | 6879<br>(6879, 6880) |
| **Annotated B-scans per Image Stack [n]** | 145<br>(140, 152) | 512<br>(512, 512) |
| **SNR<sub>B-scan</sub> [dB]** | 42.0<br>(40.2, 44.1) | 36.1<br>(34.4, 37.7) |
| **Peak B-scan Intensity [RAW value × 10<sup>6</sup>]** | 1037.4<br>(756.1, 1470.4) | 954.2<br>(737.7, 1610.7) |
| **Vessel Intensity [RAW value × 10<sup>6</sup>]** | 7.9<br>(2.4, 61.8) | 49.6<br>(19.9, 129.3) |
| **SNR<sub>Vessel</sub> / Peak SNR<sub>Vessel</sub> [%]** | 2.1<br>(1.5, 3.3) | 2.5<br>(1.7, 3.2) |
| **Average Velocity [a.u.]** | 14<br>(10, 17) | 48<br>(41, 58) |
| **Minimal Velocity [a.u.]** | 5.8<br>(5.6, 5.9) | 24.1<br>(23.6, 24.5) |
| **Maximal Velocity [a.u.]** | 22<br>(15, 29) | 73<br>(59, 92) |

Results are presented as median (IQR) of the average values per image stack
n: number of time-resolved image stacks; SNR: signal-to-noise ratio; SNR<sub>B-scan</sub>: SNR in the complete B-scan; SNR<sub>Vessel</sub>: Average SNR in the vessel subareas of the B-Scan; Peak SNR<sub>Vessel</sub>: maximum SNR on a pixel level in the complete vessel subarea, corresponding to the reference SNR for velocity calculations; RAW value: value extracted from the raw .vol OCT files; a.u.: arbitrary unit; IQR: interquartile range.

The matching of the OCT and the ECG device is visualised in Figure 2 as a screenshot. The corresponding recording can be found in Supplementary Video 1. It shows the processed data, including the registered time-resolved OCT B-scans, the corresponding ECG trace and the acquisition position of the B-scan on the SLO image. The B-scan size in pixels, the FOV and the conversion to real size were extracted from the metadata of the .vol files. The ECG data is presented with the time elapsed since the last R-Peak as well as the percentage of the current cardiac cycle completed. Minor eye movements during the acquisition occurred. The registration process can compensate for motion in the anterior-posterior and fast-scanning directions. This can be identified through the black boxes at the border of the images. However, movements that were non-collinear with the fast-scanning direction could not be compensated as eye tracking was disabled.

**Fig. 2.**
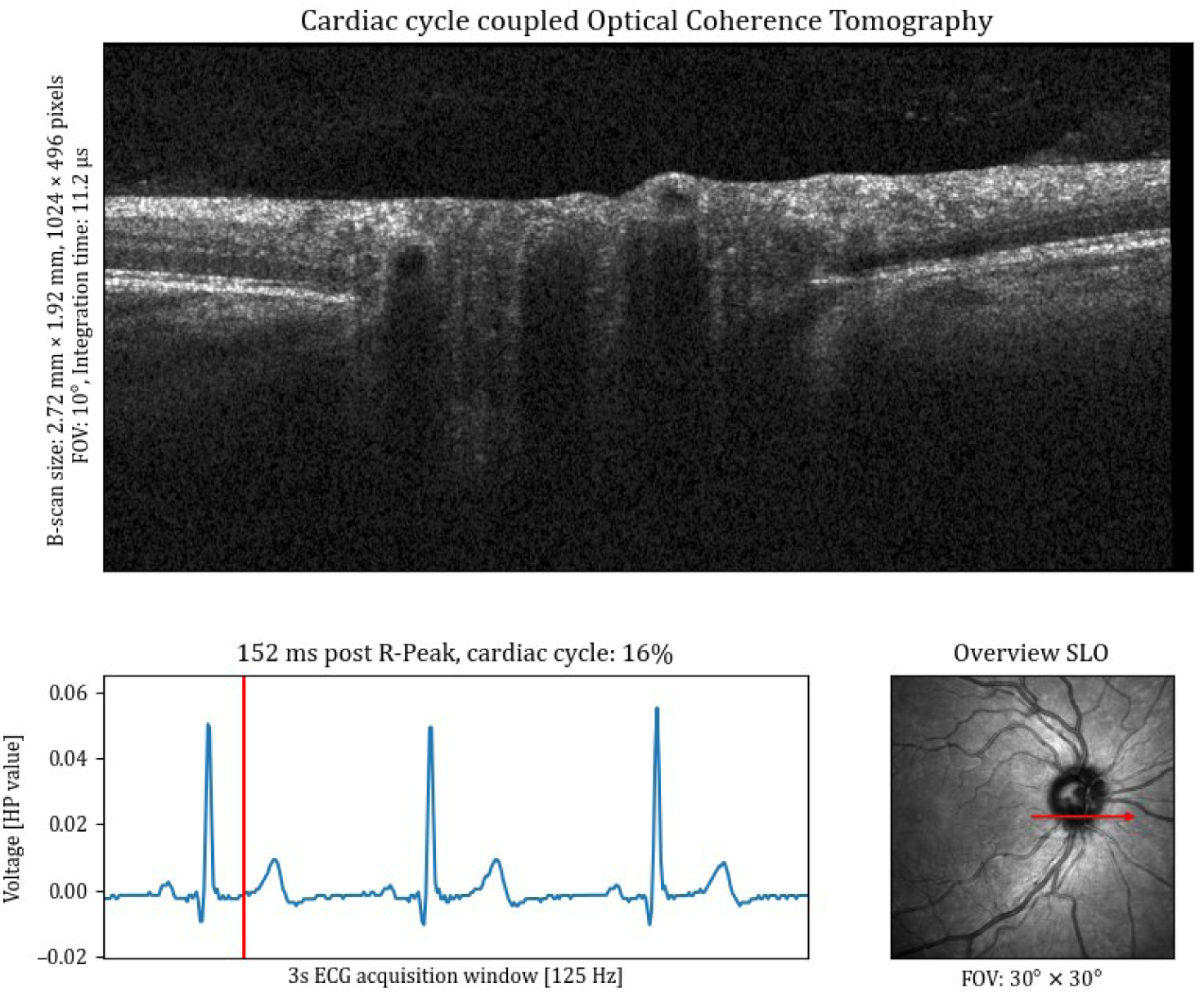
Overview of a ECG-coupled dynamic time-resolved OCT acquisition. The upper half of the image shows the B-scan that corresponds to the specific moment in the cardiac cycle. The cardiac cycle is indicated in the bottom left, where the red vertical line indicates the current moment. The red arrow in the image on the bottom right shows the acquisition location of the B-scan. FOV: field of view. SLO: Scanning laser ophthalmoscopy, ECG: electroctrocardiogram, HP value: Healthy Pi ECG voltage values.

### Correlation between the cardiac cycle and retinal pulse waves

Supplementary Video 1 shows the clear pulsatile intensity changes at the centre of the arterioles at the inferior part of the optic nerve head. These pulsatile intensity changes due to fringe washout reflect changes in the flow speed within the retinal vessels ^11^. The pulsations in the calculated blood flow profiles corresponded int in frequency to the heart rate. The envelope function of the maximal calculated flow velocity allowed to identify the local sharpest rises of the pulse wave. The sharpest rises of the pulse wave were used to determine the arrival of the pulse wave at the retina. The plotting of the sharpest rise and the R-Peaks revealed that both peaks occured with relative constant lags.

The example OCT subarea SNR, the calculated axial flow velocity profiles, the velocity gradients with the sharpest rise, and the ECG trace with the R-Peaks are visualised in Figure 3 for the right artery (A1) of the Supplementary Video 1. Supplementary Video 2 show another example of ECG-coupled OCT at the optic nerve head. The ocular blood flow pulsations match in frequency with the heart beat.

**Fig. 3.**
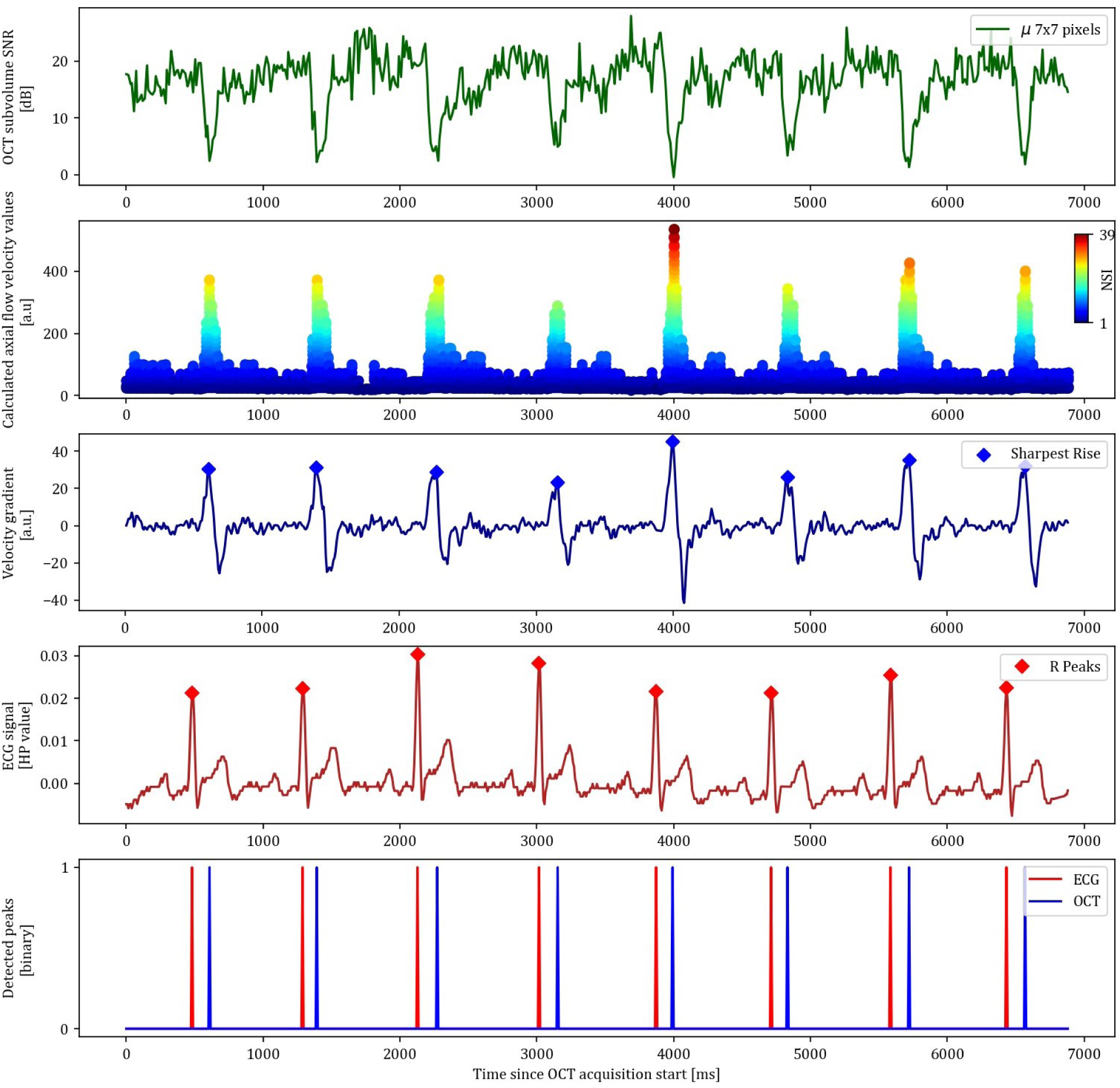
Peak signal extraction from OCT and ECG data. The time since OCT acquisition start on the x-axis is the same for all subfigures. The first plot shows the average signal-to-noise ratio (SNR) in the extracted subareas (μ 7×7 pixels) over time. The second plot shows the calculated axial flow velocity values based on fringe washout, where the NSI represents the numerical solution index. The third plot shows the velocity gradient of the envelope function of the second subplot, where the sharpest rise is automatically detected. The fourth plot shows the ECG signal from the HealthyPi (HP) with detected R-Peaks. The detected peaks from both ECG and OCT are then plotted on top of each other in the fifth plot.

The calculated axial flow profiles are represented as a scatter plot, where each B-scan is represented as a pillar of scatters on the x-axis. Each numerical solution to the SNR_Drop_ equation is represented as a scatter, where the index of the numerical solution is indicated in the colormap. The velocity gradient is calculated for the envelope function of the maximal calculated flow velocity. The sharpest rises are hence depicted as the local maxima of the first derivative of this envelope function. The “EngZee” algorithm provided reliable detection of the R-Peaks in the ECG trace.

The timepoints of these signals were encoded as binary information with Gaussian smoothing to allow for uncertainty in the signal detection. The cross-correlation of both signals enabled to determine the most probable delay for the HRT. The ECG data was padded 10 seconds before the start and 1 second after the end of the OCT signal. The normalised cross-correlation of both signals showed the expected intermittent spikes where the argmax of the signal corresponded to the most probable peak. Figure 4 visualises the normalised cross-correlation of both signals, showing the most probable offset of the ECG to the OCT at -136 ms, which corresponds to a HRT of 136 ms. These findings showed that the R-Peaks and the sharpest rises of the pulse waves correlated well.

**Fig. 4.**
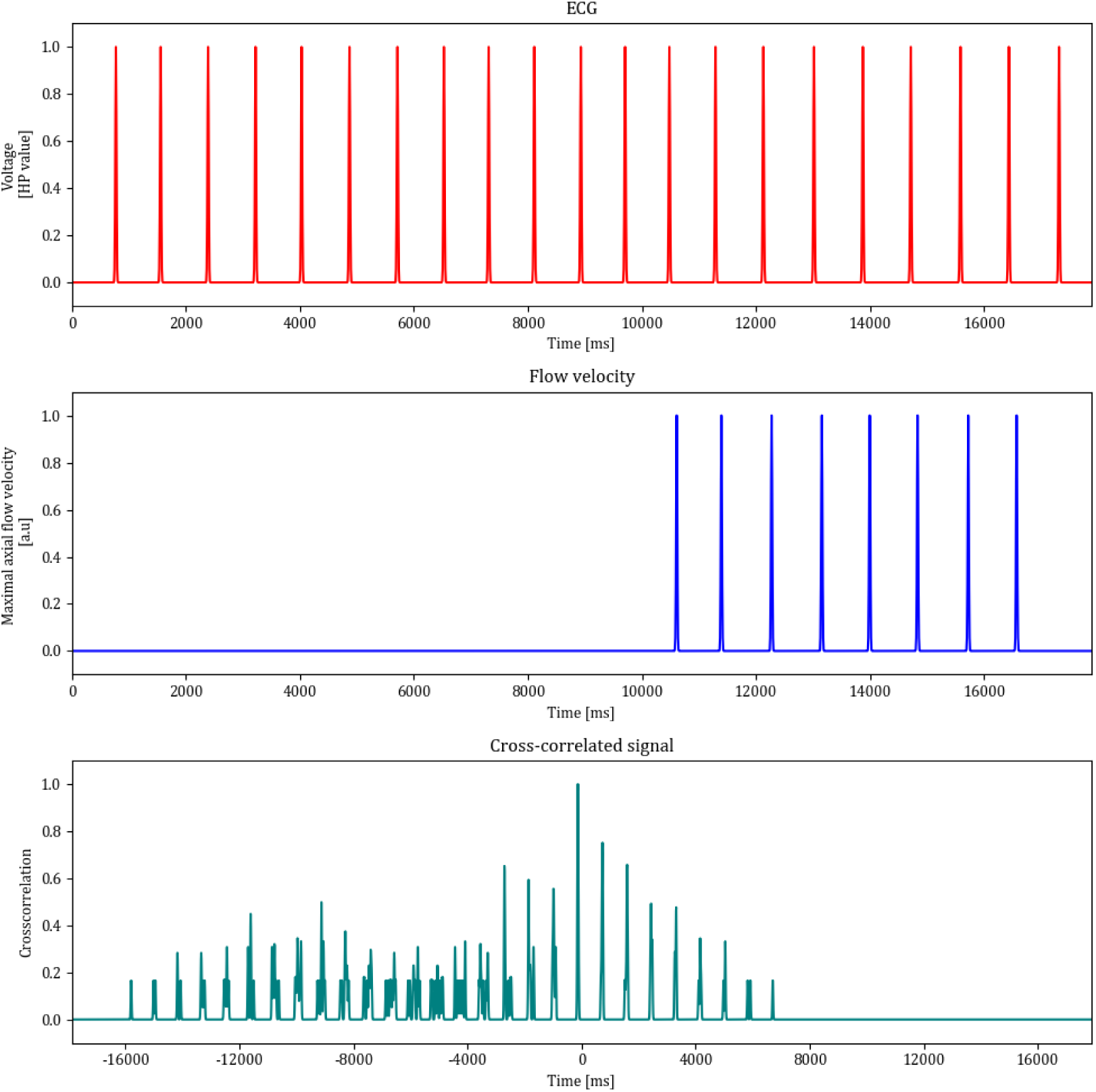
Signal analysis for the determination of the heart-retina time (HRT). The top row shows the R-Peaks from the ECG signal, the middle row shows the times of the sharpest rise in the OCT flow velocity signal and the bottom row shows the normalised cross-correlation of both signals. The ECG signal has been extended for 10s before the start and 1s after the end of the OCT signal. The highest peak in the normalised cross-correlation represents the most probable offset of the ECG towards the OCT signal, representing the additive inverse of the HRT.

### The heart-retina time as a potential biomarker

The first step to calculate the HRT was to find out how many pulse waves are on the way from the heart to the eye. As seen in Figure 4, the cross-correlation signals are repetitive peaks that occur due to the pulsatile nature of the velocity peaks. The argmax of this function provided the most probable offset for each of the 90 annotated vessels. Figure 5a shows a clearly distinguishable mode of the cross-correlation histogram at the bin of -150 to -100 ms. This indicates that the blood column between the heart and the retina is propagated within one heart cycle.

**Fig. 5.**
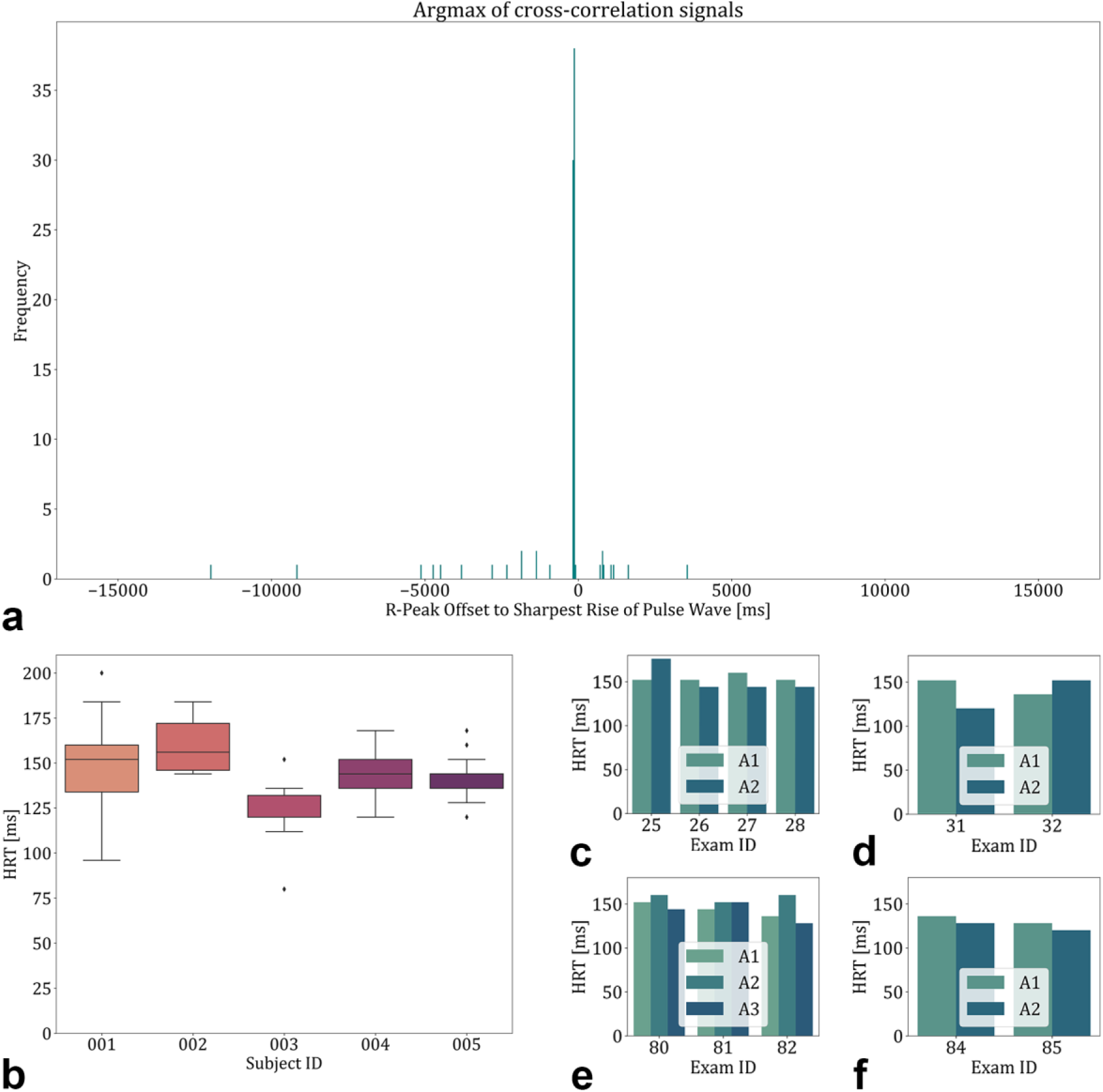
Heart-retina time (HRT) results overview. **a** Histogram plot with a bin width of 50ms for the argmax of the cross-correlation signals showing that the most probable delay between the R-Peak and the arrival of the pulse is within one cardiac cycle. **b** HRTs of the five included subjects showing good intrasubject and inter-subject reproducibility. **c-f** HRTs of different acquisitions within the same vessels.

The HRT was then calculated for each of the acquisition time series with the argmax of the cross-correlation signal within one heartbeat, the results further apart being identified as outliers. The HRT is shown as the additive inverse of the calculated offset. The HRT showed good intersubject and intrasubject reproducibility. The HRT of all acquisitions was 144 ± 19 ms (mean ± SD). A visualisation of the measurements for each subject can be found in Figure 4b. The intersubject CoV was 0.09, and the intrasubject CoV was 0.11 ± 0.03.

Figure 5c-f shows the results of several acquisitions where more than one vessel was annotated. Each subplot represents one acquisition location and each Exam ID represents one time series. The intra-exam CoV in case more than one vessel was annotated was 0.07 ± 0.04. The intra-vessel CoV in case the HRT was calculated in more than one exam was 0.09 ± 0.05. Supplementary Video 3 shows a representation of an acquisition where three arterioles were annotated and evaluated to calculate the HRT. The video visually shows the synchronous arrival of the pulse waves in all three annotated arterioles. In Figure 6, the available information from an ECG-coupled time-resolved dynamic OCT with three annotated arterioles is shown.

**Fig. 6.**
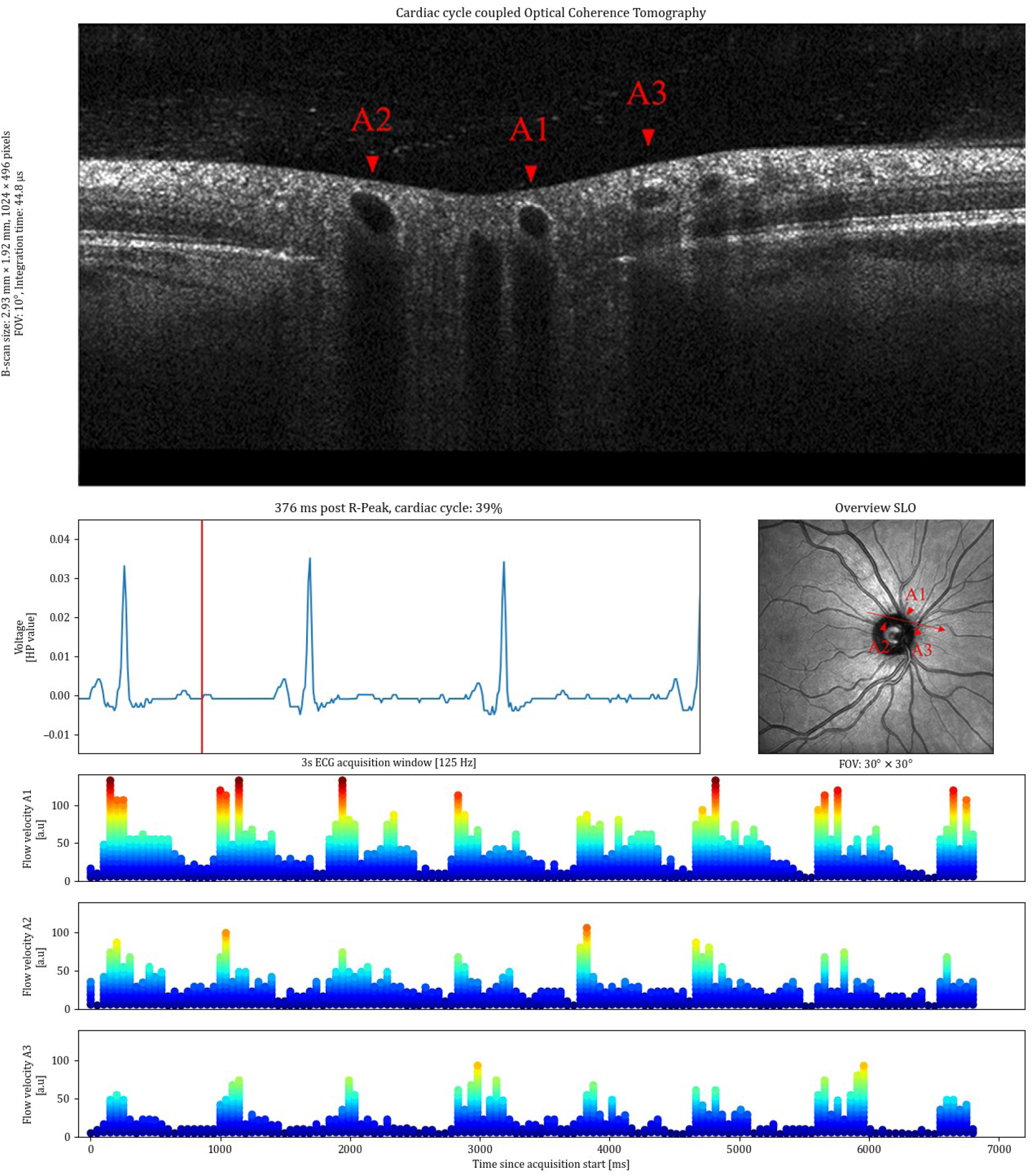
Overview of the information available in ECG-coupled dynamic time-resolved OCT, showing an acquisition with blood flow profiles from three annotated arterioles (A1, A2, A3). The identification of the arterioles in the OCT was made on the corresponding SLO image. The blood flow profiles show synchronous pulsatility, corresponding in frequency with the cardiac cycle over the synchronous acquisition of 7 seconds. The analysis of the delay allows to calculate the HRT for all of the annotated vessels

## Discussion

In this study, we developed a method to link the cardiac cycle to the blood flow pulsations in the retina. We connected a commercially available OCT device to an open-source ECG and merged the acquisitions via synchronised timestamps. Pulsatility was found in the OCT flow velocity profiles, and the sharpest rises of the pulse waves were identified. These peaks were cross-correlated with the R-Peaks from the ECG. We found that the blood column between the heart and the retina propagates within one cardiac cycle. Via an analysis of the lag between the two signals, we can calculate the HRT. We propose this HRT as a potential new biomarker for the analysis of cardiovascular health.

Cardiovascular diseases, including hypertension, frequently manifest in both the heart and the eye ^1^. Conditions like hypertensive retinopathy, which can result in vision loss, are examples of this interconnectedness ^27^. Many ideas have been developed to identify systemic diseases from the eye by gaining information about the vascular state ^28,29^. Few studies have been conducted to link the heart and the eye via ECG to assess the pulsatile vessel calibre on fundus photographs or with ultrasound to assess the larger carotid or ophthalmic arteries in synchronicity with the heart ^30–33^. To the best of our knowledge, we present the first link of an ECG with OCT to assess retinal blood flow profiles.

With this method, we showed that the blood flow propagates towards the retina within one cardiac cycle. This finding might be counterintuitive initially as the arm-retina time from fundus fluorescein angiography (FFA) is in the range of 7-15 seconds ^34^. In FFA, a dye bolus is injected into the arm of the patient and then propagated towards the peripheral circulation. The fluorescein is mainly bound to human serum proteins after injection ^35,36^. For the FFA arrival time, the dye hence travels at the same speed as the particles in the blood. In our study using ECG-coupled OCT analysis, we observed significantly shorter delay times for the HRT. This result can be described similarly to a hose filled with water: when the water pump is activated, water begins to flow out of the hose almost instantly, rather than requiring the water to traverse the entire length of the hose. When the pump is restarted, water starts almost immediately to flow out of the hose opening, without the water having to travel all the way through the hose. Hence, we hypothesise that we rather measure the propagation of the blood column instead of the propagation speed of single blood particles.

The assessment of blood flow in conjunction with the heart has been explored in several other fields, with a particular focus on the pulse transit time ^37–39^. The pulse transit time measures the time taken for blood to travel between two locations in the body. The transit time for the blood to travel from the heart, as measured from the R-Peak in the ECG, to the carotid artery has been measured at 100 ± 11 ms ^32^. The pulse transit times for the blood to arrive from the heart to the ear, finger, and toe have been measured at 126, 269, and 266 ms, respectively ^40^. The measured HRT of 144 ± 19 ms is well compatible with these previous findings.

Pulse transit times, in general, have been shown to correlate well with blood pressure and have often been suggested as an additional biomarker, extending or even substituting cuff-based blood pressure measurements ^38,39,41^. The pulse wave velocity can be calculated by dividing the distance between the two measurements by the pulse transit time ^42^. Shorter pulse transit times hence correspond to faster pulse wave velocities, and faster pulse wave velocities are found in stiffer arteries ^43^. The analysis of pulse wave velocity can not only be used to estimate the blood pressure but also to assess target organ damage ^44^. Faster pulse wave velocities have been found to be associated with the risk of stroke and to be predictive of cerebrovascular events ^45^. As the retina shares a significant proportion of the perfusing vessels with the rest of the brain, the HRT has the potential to lead to a better understanding of the pulse wave propagation towards arterioles perfusing the central nervous system.

With the proposed HRT, we aim to contribute to the field of pulse arrival times. The HRT in this first feasibility study showed good intra-subject, inter-subject, intra-exam and intra-vessel reproducibility. The intra-subject, inter-subject and intra-vessel variabilities were in a very similar range. We estimate these variations to be attributable to noise and natural variability, which could be similar to the heart-rate variability. The heart-rate variability can be used for an objective assessment of stress^46^. The intra-exam CoV showed the lowest variability, however, it was still existent.

This variability between different vessels in the same examination comes through different pulse wave forms in the OCT flow profiles, as the identified R-Peaks are the same for all within the same examination. The further exploration of HRT variability, particularly its relationship with heart-rate variability, could help differentiate between random fluctuations and inherent variability. In further investigations of HRT variability, valuable information about the propagation of blood flow could be gained.

The study has limitations of different origins. One is the small number of subjects included in the study. This was partly compensated by the repetitive acquisition and will be further addressed by generating normative databases in a larger population in the future. Future work will also expand the study populations to include subjects with cardiovascular diseases to evaluate and validate the potential of the HRT as a biomarker. Another factor is the synchronisation mechanism, which consists of a calibration of the PC and ECG time series via NTP. In the future, mechanisms to use the same clock for the ECG and OCT could be investigated. In this work, we were limited by the used devices, which did not allow for a direct access of the clock of the other instrument, potentially leading to a HRT biases between different acquisitions. A further limitation is the time-consuming analysis procedure of single arteriole centres. However, the 26’100 annotated vessel centres from this study could serve as the basis for the development of automated landmark detection systems, alleviating the manual burden.

Our study’s main strength is that we developed a method with a widely used commercially available OCT device. Second, the ECG device we used is based on an open-source device, which would allow for an easy integration with different OCT devices. Hence, this means that the method can be used and reproduced in other settings, where time-resolved OCT acquisitions are feasible. Third, we propose a quantitative method to assess the cardiovascular state. Similar to other pulse wave velocity methods, we provide a specific method for the propagation from the heart to the eye. Future studies will assess the validation and clinical usability of the HRT as a biomarker for cardiovascular health.

## Conclusions

In this study, we propose a method to analyse the propagation of blood flow from the heart to the retina. We demonstrated the feasibility of ECG-coupled time-resolved OCT acquisitions and the correlation between the cardiac cycle and retinal pulse waves. The cross-correlation of the ECG and the blood flow profiles in retinal arterioles revealed the propagation of the blood column within a cardiac cycle. By analysing the lag between the two signals, the HRT can be calculated. The investigation of blood flow propagation is an important field to better understand vascular turbulences. The synchronised acquisition of ECG with dynamic OCT offers a method to explore cardiovascular and ocular health interconnections. This study establishes the foundation for utilising the HRT as a potential biomarker in evaluating systemic and ocular vascular conditions.

## Acknowledgments

This study received funding via personal grants to PV from the Swiss National Science Foundation (Grant 323530_199395), AlumniMedizin Basel and the Janggen-Pöhn Foundation. In addition, the researchers would like to thank all the participants who volunteered for this study.

## Author contributions

PV: Conceptualisation, methodology, software, validation, formal analysis, investigation, resources, data acquisition, data processing, writing (original draft preparation), writing (review and editing), visualisation and project administration.

JW: Methodology, software, validation, formal analysis, investigation, writing (review and editing).

FB: Methodology, software, validation, formal analysis, investigation, writing (review and editing).

HPNS: Methodology, validation, investigation, resources, writing (review and editing) and project administration.

PCC: Conceptualisation, methodology, software, validation, formal analysis, investigation, resources, data processing, writing (original draft preparation), writing (review and editing) and project administration.

PMM: Conceptualisation, methodology, validation, formal analysis, investigation, resources, data processing, writing (original draft preparation), writing (review and editing), visualisation and project administration.

## Ethics declarations

PV received speaker fees from Heidelberg Engineering GmbH and Bayer. HPNS is supported by the Swiss National Science Foundation (Project funding: “Developing novel outcomes for clinical trials in Stargardt disease using structure/function relationship and deep learning” #310030_201165, and National Center of Competence in Research Molecular Systems Engineering: “NCCR MSE: Molecular Systems Engineering (phase II)” #51NF40-182895, the Wellcome Trust (PINNACLE study), and the Foundation Fighting Blindness Clinical Research Institute (ProgStar study). Dr. Scholl is member of the Scientific Advisory Board of: Boehringer Ingelheim Pharma GmbH & Co; Droia NV; Eluminex Biosciences; Janssen Research & Development, LLC (Johnson & Johnson); Okuvision GmbH; ReVision Therapeutics Inc.; and Saliogen Therapeutics Inc. Dr. Scholl is a consultant of: Alnylam Pharmaceuticals Inc.; Gerson Lehrman Group Inc.; Guidepoint Global, LLC; and Tenpoint Therapeutics. Dr. Scholl is member of the Data Monitoring and Safety Board/Committee of Belite Bio (DRAGON trial, NCT05244304; LBS-008-CT02, NCT05266014), F. Hoffmann-La Roche Ltd (VELODROME trial, NCT04657289; DIAGRID trial, NCT05126966; HUTONG trial), ViGeneron (protocol number VG901-2021-A) and member of the Steering Committee of Novo Nordisk (FOCUS trial; NCT03811561).

PMM is a consultant of Roche and holds intellectual properties for machine learning at MIMO AG and VisionAI, Switzerland.

The other authors declare no conflict. Funding organisations had no influence on the design, performance or evaluation of the current study.

## Data availability

The data presented in this paper are not publicly available due to data protection regulations. Interested parties may request access to the data from the corresponding author (PV) upon reasonable request and approval from the concerned institutional review boards.

## Supplementary Material

**Supplementary Video 1**

Electrocardiogram-coupled time-resolved dynamic optical coherence tomography at the optic nerve head. The pulsatile intensity changes at the centre of the arterioles are due to fringe washout.

**Supplementary Video 2**

Electrocardiogram-coupled time-resolved dynamic optical coherence tomography of an arteriole with synchronised velocity profile and electrocardiogram trace at the optic nerve head. The ocular blood flow pulsations match in frequency with the heart beat.

**Supplementary Video 3**

Electrocardiogram-coupled time-resolved dynamic optical coherence tomography of three arterioles with synchronised velocity profiles and electrocardiogram trace at the optic nerve head. The ocular blood flow pulsations match in frequency. The arrival time can be calculated and is denoted as the heart-retina time.

